# GLP-1/GIP Uptake, Indication, and Access Pathways Among US Adults in the Understanding America Study

**DOI:** 10.64898/2026.08.28.26361368

**Authors:** Ritika Chaturvedi, Tadeja Gracner, Francisco Perez-Arce, Sze-chuan Suen, Jing Jin, Bart Orriens, Rosalie Liccardo Pacula, Alison Sexton Ward, Rebecca Haile, Arie Kapteyn

## Abstract

**Importance:** Evidence on GLP-1/GIP therapies is largely derived from trials enrolling selected populations or medical records that miss utilization outside healthcare channels. No nationally representative cohort has characterized real-world uptake, indications, and access.

**Objective:** To characterize GLP-1/GIP prevalence, indication, clinical profile, and access.

**Design:** Prospective cohort study with three GLP-1/GIP surveillance waves (March 2024, December 2024, October 2025).

**Setting:** The Understanding America Study, an address-based, nationally representative panel of approximately 15,000 US adults aged 18+ years initiated in 2014.

**Participants:** UAS participants responding to at least one surveillance wave (n=9150).

**Exposures:** GLP-1/GIP use status (never vs any use, comprising current and former use), self-reported primary indication (diabetes, weight loss, or other), and access pathway (traditional vs non-traditional).

**Main Outcomes and Measures:** Survey-weighted prevalence of GLP-1/GIP use, overall and by indication and access pathway; sociodemographic, cardiometabolic, treatment, and access characteristics; and smartwatch-derived resting heart rate, heart rate variability, maximum activity heart rate, step count, and sleep duration and variability.

**Results:** Among n=9150 adults (1274 with any use; 60.9% female; median age 53 years), weighted prevalence increased 46%, from 8.2% (March 2024) to 12.0% (October 2025) representing 32 million. Weight-loss indications grew, reaching nearly half of use (4.1% to 5.6%); diabetes-indicated use was stable (5.3% to 5.4%). Users carried high cardiometabolic burden (obesity, 68.2%; diabetes, 53.6%) but diverged by indication: diabetes-indicated users were older (median, 59 vs 49 years), whereas weight-loss-indicated users were more often female (69.9% vs 51.3%) and healthier. One in three users (~9 million) had non-traditional access, especially in weight-loss-indicated users, of whom 33% had no conventional prescription; 41% used compounding, online, or foreign pharmacies; and, 43% lacked coverage. Non-traditional users were five times as likely to report an unlisted, likely compounded formulation (19.8% vs 4.1%). All p<0.05.

**Conclusions and Relevance:** Real-world GLP-1/GIP use has grown rapidly and diversified substantially in indication, access, and population profile. One in 3 users obtained treatment through nontraditional channels largely invisible to claims data, raising long-term safety, efficacy, and coverage questions. GLIMMER provides a public, nationally representative longitudinal evidence base for future payer and provider decisions.

**KEY POINTS:** *Question:* Who is using glucagon-like peptide-1 receptor agonists or glucose-dependent insulinotropic polypeptides (GLP-1/GIP) therapies in the US, how has use changed since 2024, and how are these medications obtained?

*Findings:* In this cohort study of 9150 US adults, GLP-1/GIP use rose significantly from 8.2% to 12.0% (March 2024–October 2025), driven by weight loss; those treated for diabetes were a decade older, lower-income, and in poorer health than those treated for weight loss. Overall, 1 in 3 users obtained treatment outside conventional prescribing and dispensing channels.

*Meaning:* A large and growing share of GLP-1/GIP exposure occurs outside the channels visible to claims-based surveillance.

## INTRODUCTION

Glucagon-like peptide-1 receptor agonists and glucose-dependent insulinotropic polypeptides (GLP-1/GIPs) have transformed cardiometabolic treatment, with randomized trials demonstrating substantial improvement in glycemic control, weight, and cardiovascular outcomes.^1-6^ However, real-world generalizability of trial evidence is limited.^7^ Trials systematically exclude younger, healthier adults who may experience attenuated benefits and older adults at heightened risk of adverse events and drug interactions.^8^ Administration is closely supervised, eliminating real-world variation in adherence and dose escalation. Follow-up rarely exceeds 1-3 years despite lifelong intended treatment, precluding assessment of delayed benefits against harms including musculoskeletal decline, malnutrition, maladaptive behaviors, or disordered eating.

Real-world observational studies from electronic health records and administrative claims offer improved external validity,^9^ but are limited by fragmented measurement, loss to follow-up, and systematic underrepresentation of individuals facing healthcare barriers.^10,11^ Moreover, restrictive formularies, particularly for obesity,^12,13^ may drive patients toward cash-pay or direct-to-consumer access (telehealth platforms, medical spas, compounding pharmacies) none of which generate claims and are outside clinical surveillance. National surveys extend population coverage but typically lack longitudinal tracking from pre-treatment baselines,^14,15^ objective measurement,^16,17^ or the survey instrument agility required to capture a rapidly changing therapeutic and regulatory landscape.^18^ Across sources, publicly available individual-level data permitting independent replication are almost entirely absent.^19^

Both the National Institutes of Health (NIH)^20^ and the Congressional Budget Office (CBO)^21^ have called for rigorous, public, population-representative evidence on the safety and efficacy of real-world GLP-1/GIP treatment. Meeting that need requires an observational cohort that enrolls adults independent of insurance status or access channel, observes them from pre-treatment baselines forward, and releases data publicly through findable, accessible, interoperable, and reusable (FAIR) standards. We established GLIMMER (GLP-1/GIP Influence on Metabolism and Mind Examined in the Realworld, Realtime), a nationally-representative GLP-1/GIP population health study embedded in the Understanding America Study (UAS).^22^

Here we report the first 18 months of surveillance, characterizing GLP-1/GIP uptake, indication, and clinical profile, and quantifying the share of treatment obtained through traditional as well as outside conventional prescribing and dispensing channels.

## METHODS

### Study Design and Cohort

GLIMMER is an ongoing, prospective cohort study designed to fill gaps in existing GLP-1/GIP evidence infrastructure (Table 1). GLIMMER is embedded in the Understanding America Study (UAS),^23^ a nationally representative internet panel of approximately 15,000 US adults aged 18 years or older, established in 2014 (Fig 1A). UAS recruits through address-based sampling and provides hardware (smartphone, Internet, digital sensors) to participants lacking access, reducing digital-access bias.^24,25^ The panel fields Spanish-language instruments, uses periodic replenishment cohorts, and applies national poststratification weights, and closely matches national prevalence for key GLP-1/GIP indications (diabetes, 13.5%; obesity, 45%; cardiovascular disease, 8%). Cumulative response exceeds 75% and annual attrition is below 8%.

**Table 1:** Comparing UAS-GUMMER data attributes to GLP-1/GIP-related clinical trials and observational studies.

| Data attributes for GLP-1/GIP evidence generation |  | UAS-GLIMMER | Trials / Meta-Analyses | Observational Studies |
| --- | --- | --- | --- | --- |
| Population | Nationally-representative | Yes | No | No |
|  | Real-world | Yes | No | Partial (e.g., limited to observations within healthcare systems) |
|  | Inclusive of diverse GLP-1/GIP patients across healthcare access, treatment patterns | Yes | No | Partial (limited to certain insurance groups and healthcare systems) |
| Follow-up | Pre-treatment baselines | Yes | N/A (controlled) | Partial |
|  | Near-real-time surveillance | Yes | N/A (controlled) | No |
|  | Long-term, life-course follow-up | Yes (as follow-up accrues) | No (1–3 years typical) | Partial (limited to retained samples) |
| Outcomes | Treatment engagement, persistence, and patterns | Yes | No (controlled, protocol-driven) | Partial (e.g., prescription fills) |
|  | Treatment access and barriers | Yes | No | No |
|  | Clinical outcomes (objective) | Partial (validated instruments, wearable biometrics, linked claims) | Partial (limited to trial endpoints) | Partial (limited to healthcare visits) |
|  | Holistic behavioral and physical/mental/cognitive health outcome | Yes | Partial (limited to trial endpoints) | Partial (limited to healthcare visits) |
|  | Comprehensive adverse effects and maladaptive behaviors | Yes | Partial (1–3 years typical) | No |
|  | Socioeconomic outcomes and context | Yes | No | Partial (e.g., claims-related costs) |
| Analysis Potential | Identification of responder subgroups across biological, behavioral, psychosocial dimensions | Yes | Partial (predetermined stratification, e.g., diabetes v. obesity) | Partial (stratification based on observables in healthcare visits) |
|  | Causal inference | Yes (quasi-experimental research designs) | Yes | No |
|  | Individual- and population-level forecasting | Yes (microsimulation, digital twins, AI/ML classification and prediction) | Partial (short-term, trial-based extrapolations) | Partial (e.g., claims-based forecasts) |
|  | Public availability of individual-level data | Yes | No (aggregate reporting) | Partial |

Notes: Trials include STEP, SURMOUNT, ELIXA, LEADER, SUSTAIN, EXCEL, REWIND, PIONEER, AMPLITUDE, SURPASS. Observational studies include claims and electronic health record analyses, plus RAND and KFF national surveys.

**Figure 1.**
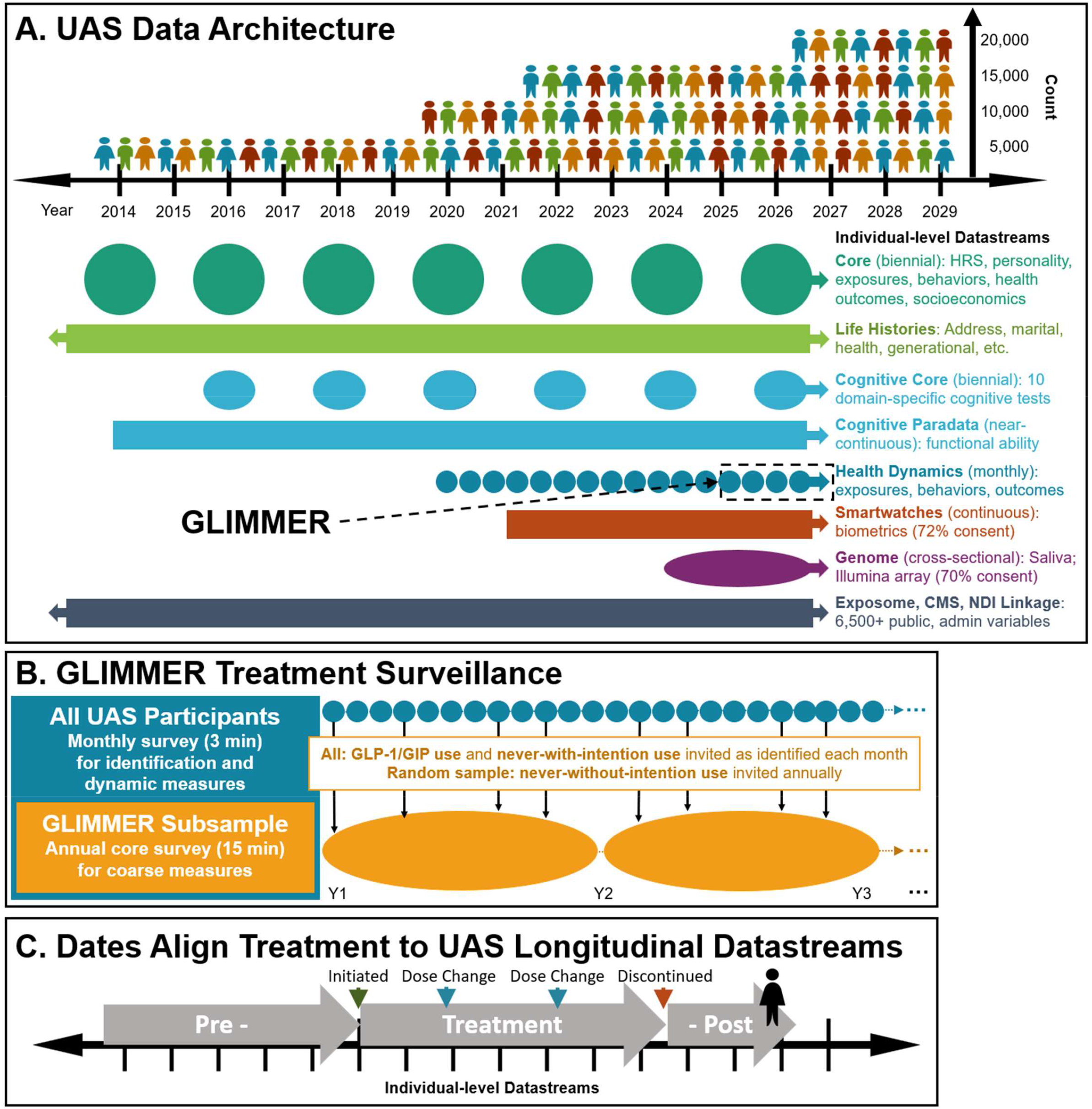
Schematic representation of GLIMMER’S design. **(A)** UAS cohort growth and deep phenotyping data architecture. **(B)** GUMMER treatment surveillance design leveraging UAS monthly surveys. **(C)** Timestamped milestones from treatment surveillance protocol allow alignment to UAS’s comprehensive longitudinal Datastreams in (A).

GLIMMER was approved by the Biomedical Research Alliance of New York Institutional Review Board (BRANY: 22-030-1044) and follows STROBE guidelines.

### Surveillance protocol

All UAS panel members were invited to monthly health tracking surveys containing a brief (<3-minute) GLP-1/GIP module (Fig 1B, with more detail in eFig 1).^26,27^ Respondents report current treatment status each month permitting near-real-time observation of transitions among 1) current use, 2) former use, 3) never use with intention to use, and 4) never use without intention, followed by a short battery of dynamic exposure and outcome measures relevant to each subgroup. A longer core instrument (~15 minutes) is administered on first identification of any use or intended use (groups 1-3) and repeated annually thereafter, together with a randomly selected and annually replenished sample of never-users without intention.^28^ Participants report dates for all treatment milestones, permitting temporal alignment with other UAS data streams. Instruments are revised within days in response to market and regulatory change ([e.g., Caremark’s removal of Zepbound preferred status (July 2025); approval of oral Wegovy and Foundayo (Q1 2026)].

Self-reported items (eTable 1) are split across instruments for temporal relevance, maximizing depth and breadth of elicited information while minimizing recall biases and participant burden. GLIMMER surveys include questions on (1) actual/intended treatment; (2) engagement: initiation/discontinuation date, indication, agent, formulation, dose, administration; (3) barriers: insurance coverage, healthcare access, utilization, dispensation, supply constraints, out-of-pocket spending; (4) treatment-specific health, behavioral, and socioeconomic outcomes: weight, glycemic status, cravings, diet, substance use, relevant diagnoses (including emerging indications like sleep apnea, addiction, poly-cystic ovary syndrome, arthritis), employment (e.g., workplace productivity, absenteeism); and, (5) safety: gastrointestinal symptoms, nutritional deficiencies, sleep disturbance, fatigue, hormonal symptoms, vision loss, disordered eating, frailty and musculoskeletal decline, and cancer. Participants report occurrence dates for all treatment engagement milestones and health outcomes for alignment with other UAS data streams (Fig 1C).

GLP-1/GIP surveillance responses merge with UAS variables at the person-time level. Additional UAS cohort infrastructure, including linked smartwatch, cognitive, genomic, and administrative data, is described in detail in respective publications and the UAS website.^23-25,29,30^

### Cohort Definition

We included adults reporting GLP-1/GIP use status in any of 3 surveillance waves (March 2024; December 2024–February 2025; October–November 2025; response, 68%-80%). Prevalence estimates use all respondents at each wave. For characterization of users, the resulting sample was merged at the participant-time level with 2 longitudinal files. First, sociodemographic and health measures were drawn from the UAS Comprehensive File (v03/2026; response >80%),^30^ with each participant contributing a single observation from their most recent treatment-aligned assessment. Second, Fitbit intraday data were drawn from the UAS activity data explorer,^31^ restricted to participants with at least 1 Fitbit record between December 2024 and December 2025 (>66% of person-minutes).

### Exposures and Measures

The primary exposure was GLP-1/GIP use, classified as never use or any use, where any use comprises both current and former users. Among respondents who report when use began, we construct the prevalence of use over time within the panel. Secondary exposures were self-reported primary indication (diabetes, weight loss, or other) and access pathway. Access was classified as non-traditional if the participant reported no prescription, a prescription from an online provider or medical spa, or fills from a compounding, online, or foreign pharmacy; all other users were classified as traditional. Access measures reported individually included prescription source and dispensing channels, the two components of the access classification, along with insurance type and drug coverage. Treatment measures included agent, formulation (including unlisted formulations, interpreted as likely compounded), treatment duration (days from self-reported initiation to discontinuation or most recent assessment), discontinuation, and dose escalation. Clinical measures included overweight and obesity (body mass index ≥25 and ≥30 kg/m^2^), diabetes, hypertension, heart disease, stroke, and insulin use; health care use was captured as any physician visit in the prior 6 months. Sociodemographic measures were age, sex, educational attainment, and household income.

Smartwatch measures, derived from Fitbit intraday records among consenting participants and aggregated to person-day means, included resting heart rate (RHR; minimum heart rate per participant-day, used as a proxy for true resting heart rate), heart rate variability (HRV; root mean square of successive differences between adjacent inter-beat intervals), maximum activity heart rate (HRmax; peak heart rate during active periods), and step count. Biologically implausible observations (<3% of records) were excluded prior to aggregation, consistent with standard sensor time-series preprocessing.

### Statistical Analysis

We estimated survey-weighted prevalence (in %) of GLP-1/GIP use at each wave. Continuous variables are reported as means (with standard deviations or SDs) or medians and categorical variables as proportions, overall and by use status (any vs never use), indication (diabetes versus weight loss), and access pathway (traditional versus non-traditional access). Group differences were tested using two-sided t tests. We compared user characteristics with baseline characteristics of participants in published GLP-1/GIP trials, grouped by primary outcome (weight loss vs type 2 diabetes) and pooled with sample-size weighting. Analyses were conducted from February to June 2026 using Stata version 18.0 (StataCorp)^32^ with UAS-provided poststratification weights.^33^

## RESULTS

### Cohort

The analytic cohort comprised 9150 adults, including 1274 with any GLP-1/GIP use since 2018 and 945 never-users reporting intention to use. Most participants with any use had at least 3 years of pre-initiation observation and more than 1 year of post-initiation follow-up within UAS. A subsample of 1550 participants contributed smartwatch data, including 156 with any GLP-1/GIP use.

### Prevalence and Indication

Rapid GLP-1/GIP growth is driven by weight-loss indications. Weighed prevalence of any GLP-1/GIP use rose from 8.2% of US adults in March 2024 to 9.8% in December 2024 and 12.0% in October 2025 — approximately 32 million adults, a 46% relative increase over 18 months (Fig 2A). Those who initiated before 2024 did so predominantly for diabetes, whereas weight loss dominates among more recent initiators. By October 2025, nearly half of any use was primarily indicated for weight loss, growing 36% from December 2024 (from 4.1% to 5.6% of US adults), while diabetes-indicated use was flat (5.3% to 5.4%). Semaglutide retained the largest share but declined from 54% to 50% of use (Ozempic, 41.0%; Wegovy, 9.1%), while tirzepatide grew from 22% to 30% (Mounjaro, 20.1%; Zepbound, 10.3%). New tirzepatide initiation exceeded semaglutide initiation throughout, at a similar relative rate before and after 2025 despite the introduction of Zepbound (Fig 2B).

**Figure 2.**
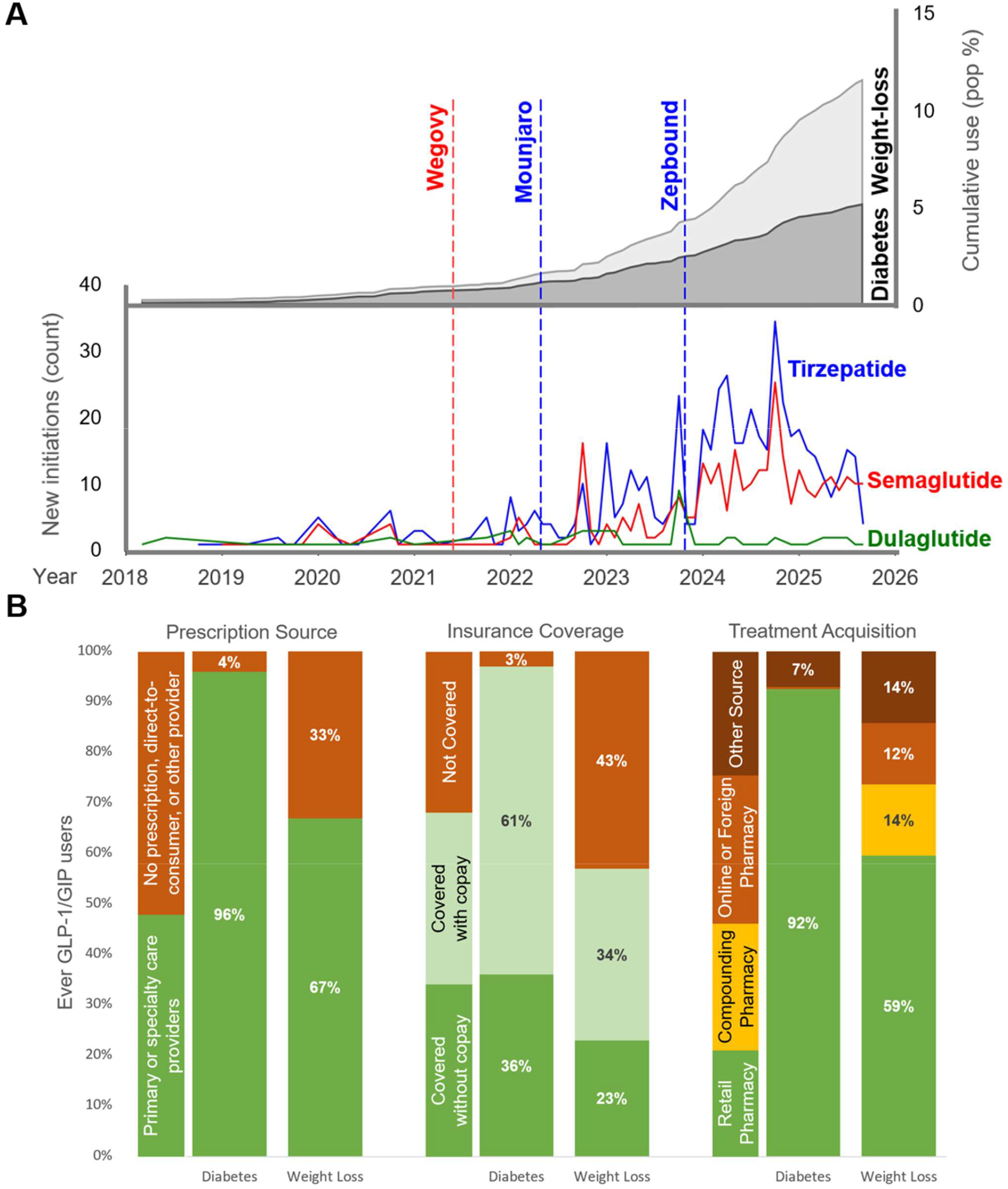
**(A)** GLP-1/GIP treatment uptake since 2018. Top: Cumulative population prevalence of “any use” for diabetes (red) and weight loss (blue) indications. Bottom: New initiations for dulaglutide (green), semaglutide (red), and tirzepatide (blue). Vertical lines = FDA approvals for specific agents. **(B)** Access and utilization landscape for US adults with any GLP-1/GIP use by primary indication.

### Clinical Profile

Adults with any GLP-1/GIP use carry greater cardiometabolic risk. Adults with any use were older than never-users (median, 53 vs 48 years), more often female (60.9% vs 49.8%), and carried substantial cardiometabolic burden (diabetes, 53.6%; obesity, 68.2%; hypertension, 61.3%) (Table 2). Consistent with greater cardiometabolic burden, users in the smartwatch subsample had higher resting heart rate (61.0 vs 57.5 beats per minute), lower heart rate variability (27.6 vs 30.6 ms), lower maximum activity heart rate (120.4 vs 123.6 beats per minute), approximately 1400 fewer daily steps (3420 vs 4858) than never-users, and 20 minutes less sleep per night (300 vs. 321 min).

**Table 2:** Weighted population estimates of GLP-1-GIP prevalence and user characteristics, overall, by diabetes vs weight loss indications, and by traditional and non-traditional access.

|  | All |  |  | Any use by indication |  |  | Any use by access |  |  |
| --- | --- | --- | --- | --- | --- | --- | --- | --- | --- |
|  | Never Use | Any Use | Pval | Diabetes | Weight Loss | Pval | Traditional | Non-traditional | Pval |
| **n (% US population)** | 7,867 (88.0%) | 1,274 (12.0%) |  | 508 (5.4%) | 590 (5.6%) |  | 799 (8.5%) | 325 (3.5%) |  |
| **Demographics** |  |  |  |  |  |  |  |  |  |
| Age (median) | 48 (16.6) | 53 (13.3) | *** | 59 (11.9) | 49 (12.8) | *** | 55 (13.0) | 51 (13.2) | ** |
| Female (%) | 49.8% | 60.9% | *** | 51.3% | 69.9% | *** | 56.7% | 70.3% | ** |
| White, not Hispanic (%) | 59.6% | 62.9% |  | 64.8% | 64.4% |  | 62.6% | 71.1% | * |
| Non-white OR Hispanic (%) | 40.4% | 37.1% |  | 35.2% | 35.6% |  | 37.5% | 28.9% | * |
| Education less than Bachelor (%) | 62.3% | 69.5% | *** | 72.5% | 65.5% | † | 71.2% | 64.4% | † |
| Hh income: <\$53K (%) | 38.6% | 31.7% | *** | 39.1% | 24.0% | *** | 34.4% | 26.1% | * |
| Hh income: \$53K-\$127K (%) | 33.8% | 37.5% | † | 36.9% | 38.7% | | 37.1% | 37.7% | |
| Hh income: \$128K+ (%) | 27.6% | 30.8% | † | 24.0% | 37.3% | *** | 28.4% | 36.3% | † |
| **Health (%)** |  |  |  |  |  |  |  |  |  |
| BMI (mean; kg/m<sup>3</sup>) | 28 (6.8) | 34 (7.8) | *** | 35 (7.2) | 35 (8.1) |  | 36 (7.6) | 32 (7.5) | *** |
| Overweight (BMI = 25.0-29.9) | 34.8% | 22.0% | *** | 21.7% | 22.1% |  | 19.8% | 23.2% |  |
| Obesity (BMI ≥ 30.0) | 32.1% | 68.2% | *** | 71.5% | 68.8% |  | 74.7% | 60.1% | *** |
| Diabetes | 13.8% | 53.6% | *** | 90.8% | 23.2% | *** | 68.9% | 23.6% | *** |
| Hypertension | 36.0% | 61.3% | *** | 74.6% | 51.4% | *** | 69.8% | 46.7% | *** |
| Any heart disease | 9.7% | 17.3% | *** | 21.2% | 11.8% | ** | 19.5% | 14.2% | † |
| Stroke | 2.3% | 6.0% | ** | 9.4% | 1.6% | ** | 7.6% | 1.5% | ** |
| Insulin use | 1.9% | 18.5% | *** | 35.6% | 5.5% | *** | 25.5% | 5.8% | *** |
| No GLP-1/GIP-related diagnosis | 45.0% | 17.7% | *** | 4.6% | 26.4% | *** | 10.9% | 28.5% | *** |
| **Cognitive Tests** |  |  |  |  |  |  |  |  |  |
| Stop and Go Switch | 1.5 (0.8) | 1.6 (0.9) | *** | 1.8 (1.0) | 1.5 (0.7) | *** | 1.7 (1.0) | 1.5 (0.8) | * |
| Figure Identification | 18.7 (6.1) | 17.6 (5.9) | *** | 16.8 (5.6) | 18.8 (5.9) | *** | 17.2 (5.7) | 19.0 (6.0) | *** |
| Serial Sevens | 4.2 (1.2) | 4.2 (1.2) |  | 4.2 (1.2) | 4.2 (1.2) | † | 4.2 (1.2) | 4.2 (1.2) | † |
| Immediate and Delayed Recall | 19.0 (4.5) | 18.6 (4.2) | * | 18.4 (3.9) | 19.1 (4.4) | † | 18.5 (4.1) | 19.3 (4.3) | † |
| Verbal Analogies | 51.1 (9.2) | 50.8 (8.9) |  | 50.1 (8.8) | 52.5 (8.5) | ** | 50.6 (8.9) | 52.3 (8.5) | * |
| Number Series | 50.4 (9.4) | 48.4 (8.7) | *** | 48.3 (8.5) | 49.1 (8.7) | † | 48.1 (8.8) | 49.8 (8.3) | * |
| Picture Vocabulary | 49.3 (9.5) | 50.3 (8.8) | ** | 51.6 (8.2) | 50.3 (8.9) | † | 51.0 (8.6) | 50.3 (8.8) | † |
| Probability of Cognitive Impairment | 7.3% | 7.7% |  | 7.2% | 6.9% | † | 7.6% | 6.6% | † |
| **Biometrics (mean (SD))** |  |  |  |  |  |  |  |  |  |
| Resting Heart Rate (BPM) | 57.5 (7.7) | 61.0 (7.7) | *** | 61.5 (7.9) | 61.7 (6.6) |  | 61.2 (7.7) | 60.8 (7.0) |  |
| Max Heart Rate with Activity (BPM) | 123.6 (12.2) | 120.4 (2.0) | ** | 118.3 (11.9) | 123.0 (10.2) | * | 119.6 (12.5) | 121.1 (9.2) |  |
| Heart Rate Variability (ms) | 30.6 (14.4) | 27.6 (12.0) | ** | 26.2 (13.6) | 28.5 (10.0) |  | 28.0 (13.0) | 25.7 (9.1) |  |
| Steps (n) | 4858 (3916) | 3420 (2632) | *** | 3135 (2467) | 3481 (2514) |  | 3130 (492) | 3868 (2524) | † |
| Main sleep duration (min) | 321 (79) | 300 (91) | ** | 290 (94) | 310 (89) |  | 299 (89) | 308 (91) |  |
| Main sleep variability (min) | 123 (42) | 123 (37) |  | 124 (40) | 123 (36) |  | 124 (39) | 123 (34) |  |

Notes: \* = p<0.05, \*\* = p<0.01, \*\*\* = p<0.001, † = p<0.10. Estimates employ UAS population weights. Percentages are sample specific unless otherwise noted.

Users diverged by indication. Diabetes-indicated users were a median of 10 years older than weight-loss-indicated users (59 vs 49 years), less likely to hold a bachelor’s degree (72.5% vs 65.5% without), more likely to report household income below $53000 (38.5% vs 23.7%), and more medically complex (diabetes, 90.8% vs 23.2%; hypertension, 74.6% vs 51.4%; cardiovascular disease, 21.2% vs 11.8%; insulin use, 35.6% vs 5.4%). Weight-loss-indicated users were predominantly female (69.9% vs 51.3%), skew younger, more educated, more affluent, and healthier; 26.4% reported no currently approved or emerging GLP-1/GIP indication, compared with 4.6% of diabetes-indicated users. They also seem more active (maximum activity heart rate 123.0 vs. 118.3 bpm). All reported comparisons were significant at p<0.05.

Both groups differed from the populations in trials. Participants in seminal diabetes-indicated trials were older (mean, 63.5 years), predominantly male (37% female), and enriched for cardiovascular risk by design (96% with cardiovascular disease) (eTables 2-3). Weight-loss trial participants had higher mean body mass index than weight-loss-indicated users in our cohort and typically excluded diabetes and major cardiovascular comorbidity, whereas 23.2% of our weight-loss-indicated users had diabetes and 11.8% had cardiovascular disease. All reported comparisons were significant at p<0.05.

### Treatment Patterns

Diabetes-indicated use is more persistent and more anchored in routine clinical care. Among all adults who had ever used GLP-1/GIPs, 64.3% were continuing at their most recent assessment and 35.7% had discontinued (Table 3). Persistence differed by indication: diabetes-indicated users had longer observed treatment duration (median, 892 vs 487 days) and lower discontinuation (28.5% vs 43.0%) compared to weight-loss indicated users. Because weight-loss indications reflect more recent uptake, the difference in observed duration, however, may partly be attributable to differential follow-up time. Diabetes-indicated users were more engaged with conventional care (83.6% vs 67.5% with a physician visit in the prior 6 months) and reported marginally higher adherence to prescribed dosing (98.4% vs 94.1%), though adherence was high in both groups. Agent use was more dispersed among weight-loss-indicated users (Wegovy, 15.8%; Zepbound, 19.1%; Mounjaro, 16.2%) than among diabetes-indicated users (Ozempic, 51.4%). All reported comparisons were significant at p<0.05.

**Table 3:** Weighted population estimates of GLP-1-GIP treatment patterns, overall, by diabetes vs weight loss indications, and by traditional and non-traditional access.

|  | Any Use | By indication |  |  | By access |  |  |
| --- | --- | --- | --- | --- | --- | --- | --- |
|  |  | Diabetes | Weight Loss | Pval | Traditional | Non-traditional | Pval |
| **n (% US population)** | 1,274 (12.0%) | 508 (5.4%) | 590 (5.6%) |  | 799 (8.5%) | 325 (3.5%) |  |
| **Indication (%)** |  |  |  |  |  |  |  |
| Diabetes | 45.1% | 100.0% | 0.0% | *** | 57.3% | 14.9% | *** |
| Weight loss | 46.4% | 0.0% | 100.0% | *** | 35.0% | 74.8% | *** |
| Likely cosmetic (no approved indication) | 4.2% | 0.0% | 9.1% | *** | 1.4% | 11.1% | *** |
| Other | 8.4% | 0.0% | 0.0% | *** | 7.7% | 10.3% | † |
| **Agent (%)** |  |  |  |  |  |  |  |
| Ozempic (semaglutide) | 41.0% | 51.4% | 32.4% | *** | 42.4% | 37.4% | † |
| Wegovy (semaglutide) | 9.1% | 1.6% | 15.8% | *** | 9.7% | 7.4% | † |
| Trulicity (dulaglutide) | 6.2% | 11.0% | 1.2% | *** | 6.8% | 4.6% | † |
| Zepbound (tirzepatide) | 10.3% | 0.6% | 19.1% | *** | 9.5% | 12.3% | † |
| Mounjaro (tirzepatide) | 20.1% | 25.3% | 16.2% | ** | 22.5% | 14.2% | * |
| Likely compounded (unlisted medication) | 8.6% | 4.3% | 11.4% | *** | 4.1% | 19.8% | *** |
| All other | 13.4% | 10.1% | 15.3% | † | 9.1% | 24.1% | *** |
| **Prescription (%)** |  |  |  |  |  |  |  |
| Primary OR specialty care | 81.3% | 96.3% | 67.4% | *** | 100.0% | 34.8% | *** |
| Non-traditional prescription | 18.7% | 3.7% | 32.6% | *** | 0.0% | 65.3% | *** |
| **Dispensation (%)** |  |  |  |  |  |  |  |
| Retail pharmacy | 74.9% | 92.2% | 59.0% | *** | 100.0% | 12.4% | *** |
| Compounding pharmacy | 7.5% | 0.1% | 14.3% | *** | 0.0% | 26.1% | *** |
| Online pharmacy | 6.6% | 0.5% | 12.2% | *** | 0.0% | 22.9% | *** |
| Other (international, free-sample, friend) | 11.1% | 7.2% | 14.5% | ** | 0.0% | 38.6% | *** |
| **Treatment engagement** |  |  |  |  |  |  |  |
| Treatment duration (days) | 677 (767) | 892 (901) | 487 (552) | *** | 742 (822) | 495 (551) | ** |
| Without discontinuation (%) | 64.3% | 71.5% | 57.0% | *** | 67.7% | 53.5% | ** |
| Increased dose (%) | 65.0% | 64.0% | 66.5% | † | 67.0% | 58.2% | † |
| Indicated administration (%) | 95.9% | 98.4% | 94.1% | ** | 98.5% | 89.2% | *** |
| **Access, Utilization** |  |  |  |  |  |  |  |
| Doctor visits within prev 2 years (n) | 11 (16) | 12 (16) | 10 (16) | † | 11 (16) | 10 (16) |  |
| Doctor visits within prev 6 months (%) | 75.1% | 83.6% | 67.5% | *** | 83.3% | 54.7% | *** |
| Any health insurance (%) | 89.5% | 90.8% | 90.6% | † | 91.6% | 87.4% | † |
| Medicaid (%) | 20.1% | 22.4% | 18.1% | † | 21.6% | 16.5% | † |
| Medicare (%) | 30.9% | 43.9% | 18.6% | *** | 34.0% | 23.8% | ** |
| **Costs** |  |  |  |  |  |  |  |
| Full treatment coverage (%) | 30.2% | 36.0% | 22.7% | *** | 35.1% | 15.6% | *** |
| Coverage with co-pay (%) | 46.4% | 60.8% | 33.8% | *** | 59.0% | 8.6% | *** |
| No treatment coverage (%) | 23.4% | 3.2% | 43.5% | *** | 6.0% | 75.8% | *** |
| Average monthly OOP cost (\$) | 296 (238) | 181 (164) | 306 (231) | † | 379 (363) | 277 (188) | † |

Notes: \* = $p < 0.05$ , \*\* = $p < 0.01$ , \*\*\* = $p < 0.001$ , † = $p < 0.10$ . Estimates employ UAS population weights. Percentages are sample specific unless otherwise noted.

### Access Pathways

Adults with weight-loss indications face greater access barriers. Access differed markedly by indication (Fig 2B). Nearly all diabetes-indicated users obtained treatment conventionally: 96% held prescriptions from primary or specialty care, 92% filled at retail pharmacies, and 97% reported drug coverage. Among weight-loss-indicated users, large shares of adults reported non-traditional access: 33% reported no prescription or a prescription from an online provider or medical spa, 41% obtained fills through compounding, online, or foreign pharmacies, 11.4% reported an unlisted (likely compounded) formulation, and 43% reported no drug coverage. All reported comparisons were significant at p<0.05.

Overall, 1 in 3 adults using GLP-1/GIPs, approximately 9 million US adults, obtained treatment through non-traditional pathways. Compared with traditional-access users, they were younger (median, 51 vs 55 years), more often female (70.3% vs 56.7%), and substantially healthier, with lower body mass index (32 vs 36), diabetes (23.6% vs 68.9%), hypertension (46.7% vs 69.8%), stroke (1.5% vs 7.6%), and insulin use (5.8% vs 25.5%). They were far more likely to report no drug coverage (75.8% vs 6.0%) and no physician visit in the prior 6 months (83.3% vs 54.7%), and 5 times more likely to report an unlisted, likely compounded formulation (19.8% vs 4.1%). Notably, 11.1% reported no GLP-1/GIP-related diagnosis (e.g., obesity, diabetes) at the time of the survey, suggesting cosmetic use (vs 1.4% of traditional-access users). Despite concerns about unsupervised escalation, rates of dose escalation did not differ significantly across access pathways. All reported comparisons were significant at p<0.05.

## DISCUSSION

In the first 18 months of GLIMMER surveillance, GLP-1/GIP use among US adults rose from 8.2% to 12.0%, representing approximately 32 million adults by October 2025, consistent with contemporaneous national estimates.^17,34^ Growth was concentrated in weight-loss indications, which now account for nearly half of all use. That shift has changed who is treated: compared with diabetes-indicated users, weight-loss-indicated users are younger, more often female, more affluent, and less medically complex. Both groups differ from the populations in which the trial evidence base was generated.

These findings have direct implications for real-world surveillance as well as coverage policy. First, the real-world treated population diverges from trial populations in both directions. Diabetes-indicated trials enrolled older participants enriched for cardiovascular risk by design; the median diabetes-indicated user in our cohort is younger and considerably healthier, with 21.2% reporting cardiovascular disease.^35-38^ Whether the cardiovascular benefits demonstrated in those trials extend to this lower-risk profile, and whether earlier initiation yields greater cumulative benefit or greater cumulative harm, is not established. Conversely, weight-loss-indicated users accessing treatment through conventional channels appear less severely obese than trial participants but carry greater cardiometabolic complexity; a pattern consistent with payer requirements that condition coverage on comorbidity.^39^ This phenotype may be more vulnerable to adverse events, including musculoskeletal decline and drug interactions, that trials were not designed to detect.^40,41^

Second, drug coverage functions as a filter on what existing surveillance can observe. Coverage was near-universal among diabetes-indicated users (97%) and absent for 43% of weight-loss-indicated users. That gradient maps directly onto the access pathway: uncovered users disproportionately obtain medication through compounding pharmacies, telehealth platforms, and direct-to-consumer services that generate no claims.^39^ Claims-based data capture the covered, medically complex, diabetes-indicated user well. Yet, they systematically exclude the uncovered, weight-loss-indicated population whose patterns of use (e.g., off-label initiation, variable compounded dosing, higher discontinuation) diverge most from trial conditions and which is growing fastest.

Third, relatedly, and most consequential for surveillance, approximately 9 million US adults appear to obtain GLP-1/GIPs outside conventional clinical pathways. Three implications follow. First, compounded formulations are not subject to the bioequivalence and stability requirements applied to FDA-approved agents, introducing dose-response variability that cannot be recovered analytically downstream.^42,43^ Second, direct-to-consumer prescribing increases the likelihood of off-label or cosmetic use, consistent with the 26.4% of weight-loss-indicated users and 11.1% of nontraditional users reporting no approved indication. Finally, high observed discontinuation (42.8% among weight-loss users) diverges from the continuous-exposure assumption underlying trial extrapolations, with implications for both weight regain (projected to return to baseline within ~1.5 years among newer GLP-1 agents based on 37-study meta-analytic evidence)^44-47^ and the interpretation of cumulative-benefit estimates used in cost-effectiveness modeling, typically assuming continuous persistence.^48^ GLIMMER’s monthly surveillance with pre- and post-treatment outcomes are designed to supply exactly the real-world discontinuation and rebound trajectories these models currently lack as well as the market, regulatory, socioeconomic, and environmental factors that drive such patterns.

### Limitations

This analysis has limitations. First, exposure, indication, and access pathway are self-reported and subject to misclassification, though self-reported GLP-1/GIP use has shown good concordance with pharmacy records (Figure 2). Second, reporting of non-traditional access may be subject to social desirability bias. Third, although the cohort is followed prospectively, the comparisons reported here ascertain exposure and characteristics contemporaneously and cannot - nor are intended to - support causal interpretation. Fourth, observed treatment duration and discontinuation are subject to administrative censoring and to differential follow-up time, which is shorter for the more recently initiated weight-loss-indicated group; formal time-to-event analysis of persistence is ongoing. Fifth, the smartwatch subsample is small (156 users), and provides limited precision, particularly for comparisons stratified by indication. Finally, indication is self-assigned and may misclassify adults with dual indications.

### Conclusions

In this nationally representative cohort, GLP-1/GIP use expanded rapidly and diversified in indication, clinical profile, and access pathway over 18 months. Claims based data is insufficient at capturing the growth in their use, given the share of the population that obtain GLP-1/GIP for weight loss outside traditional medical channels. Population-level surveillance spanning indications and access pathways is necessary, not just to inform payer and provider decisions in this rapidly changing treatment and regulatory landscape, but to understand patient exposure, use and potential side-effects. Continued GLIMMER follow-up will track market and regulatory evolution and characterize real-world initiation, continuation, and discontinuation patterns not captured in claims or trial data. Data will be publicly available, permitting independent replication and further research.

## Data Availability

All data produced are available online at uasdata.usc.edu for research purposes upon signing a Data Use Agreement.

https://uasdata.usc.edu

## Acknowledgements

We are grateful to UAS participants who agreed to contribute their time and data for research. This study relies on data from the Understanding America Study, which is maintained by the Center for Economic and Social Research (CESR) at the University of Southern California and supplemented by the following grants from the National Institute on Aging, the Social Security Administration, and the National Library of Medicine: U01AG054580, U01AG077280, U01AG077280-03S1A1, R01AG083097, R01LM013237, R01AG079967.

## Author Contributions

Drs Chaturvedi and Gracner had full access to all of the data in the study and take responsibility for the integrity of the data and the accuracy of the data analysis. Drs Chaturvedi and Gracner contributed equally.

Concept and design: TG, RC

Acquisition of data: TG, RC, FPA, BO, AK

Analysis, interpretation: TG, RC, FPA, RP, ASW, RH, JJ, SS, AK

Drafting of the first manuscript: TG, RC

Critical review of the manuscript for important intellectual content: TG, RC, FPA, RP, ASW, RH, JJ, SS, AK

Statistical analysis: FPA, JJ

Obtained funding: TG, RC, AK, ASW

Administrative, technical, or material support: TG, RC, BO, RH Supervision: TG, RC

## Data Sharing Statement

UAS makes deidentified study data available to registered individuals (who sign the Data Use Agreement) for research purposes, Additional study information, such as data dictionaries, protocols, informed consent documentation, and modeling code are also available. All above information can be obtained from the UAS website at https://uasdata.usc.edu/.

## Contents

**eFigure 1.**
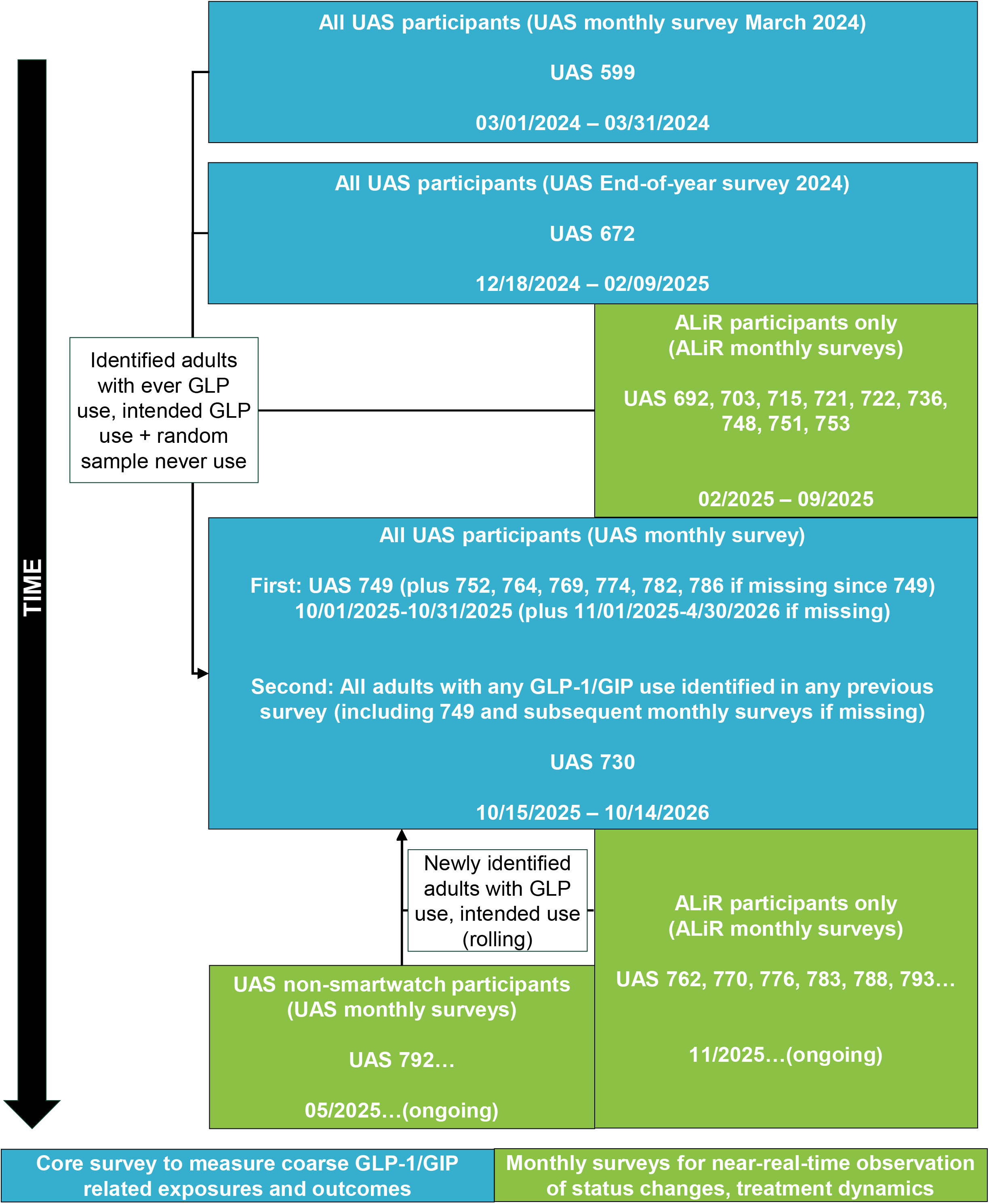
Schematic representation of individual UAS surveys used in analysis.

**eTable 1:**
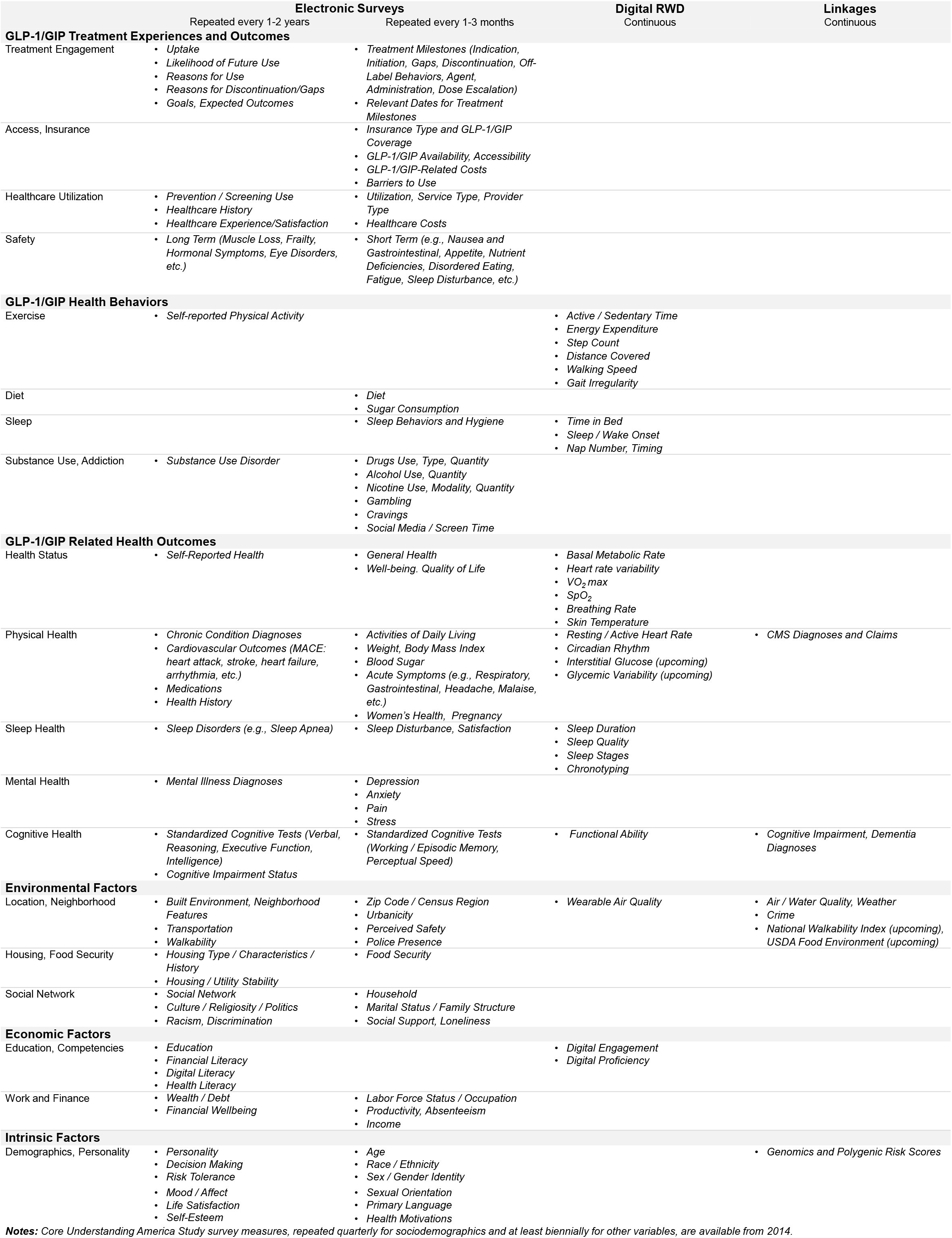
GLP-1/GIP-relevant individual data collected longitudinally in UAS (not exhaustive)

**eTable 2:**
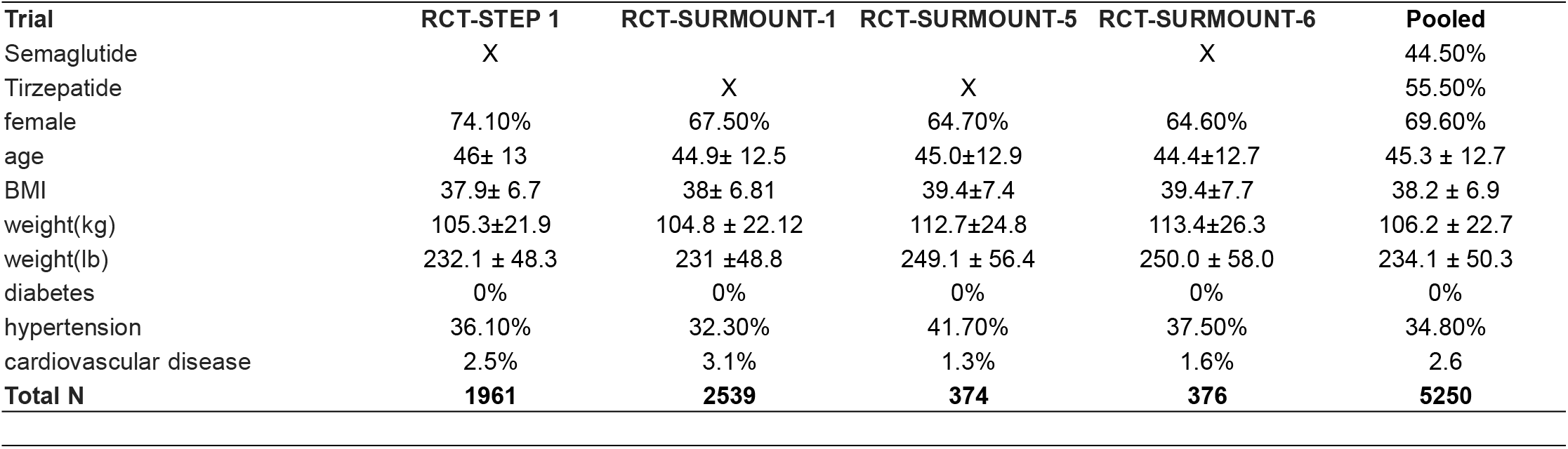
Weight-loss trials effects and pooled effects.

**eTable 3:**
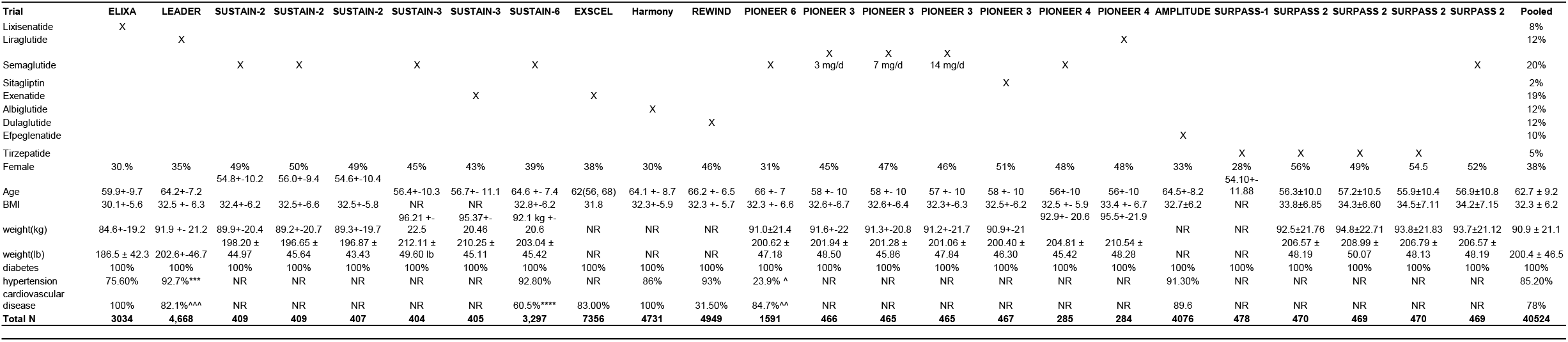
Cardiovascular outcomes trials effects and pooled effects.

